# Effects of dietary interventions focused on adding base in the management of metabolic acidosis in adults with chronic kidney disease: A protocol for systematic review and meta-analysis of randomized clinical trials

**DOI:** 10.1101/2023.04.06.23288246

**Authors:** Sepideh Mahboobi, Rebecca Mollard, Navdeep Tangri, Nicole Askin, Rasheda Rabbani, Ahmed M. Abou-Setta, Dylan Mackay

**Affiliations:** Department of Human Nutritional Sciences, Faculty of Agriculture and Food Sciences, University of Manitoba, Winnipeg, MB, Canada; Chronic Disease Innovation Center, Seven Oaks Hospital, Winnipeg, Manitoba, Canada; Department of Internal Medicine, Max Rady Faculty of Medicine, Rady Faculty of Health Sciences, University of Manitoba, Winnipeg, Manitoba, Canada; Neil John Maclean Health Sciences Library, University of Manitoba, Winnipeg, Manitoba, Canada; George and Fay Yee Centre for Healthcare Innovation, Max Rady College of Medicine, Rady Faculty of Health Sciences, University of Manitoba, Winnipeg, Manitoba, Canada; Department of Community Health Sciences, Max Rady Faculty of Medicine, Rady Faculty of Health Sciences, University of Manitoba, Winnipeg, Canada; Department of Internal Medicine, Section of Endocrinology, Max Rady Faculty of Medicine, Rady Faculty of Health Sciences, University of Manitoba, Winnipeg, Canada

**Author notes:** **Name and full address of corresponding author:** Dylan MacKay, Department of Food and Human Nutritional Sciences, Faculty of Agriculture and Food Sciences, and Department of Internal Medicine, Section of Endocrinology, Rady Faculty of Health Sciences, 196 Innovation drive, University of Manitoba, Winnipeg, MB R3T 2N2, Canada.

**Keywords:** Dietary acid load, Metabolic acidosis, Chronic kidney disease, Fruit and vegetables

## Abstract

**Background:** Metabolic acidosis is a common complication of chronic kidney disease (CKD) which can impair the function of multiple organs and accelerate CKD progression to kidney failure. The condition is usually treated with sodium bicarbonate (NaHCO3), which is an alkali salt. Although effective, alkali therapy has numerous side effects including gastric discomfort and bloating, with many people having difficulty tolerating higher doses. Research has shown that base producing fruit and vegetables may have the ability to increase serum bicarbonate concentrations similar to what is achieved with alkali therapy, but also provides added benefits. This systematic review aims to identify, critically-appraise and meta-analyze findings from randomized clinical trials (RCTs) comparing the effects of dietary interventions, including base producing fruit and vegetables, on serum bicarbonate concentrations as well as other factors related to kidney function in adult patients with CKD.

**Methods:** RCTs (in adult participants (18 years of age or older), with CKD will be included in the study. Studies will be excluded if participants are undergoing dialysis or have chronic obstructive pulmonary disease (COPD) requiring oxygen therapy. The interventions of interest are any dietary intervention aimed at manipulating dietary acid load, compared with usual care, no treatment or placebo. Our primary outcome measure will be changes in serum bicarbonate concentration, while other parameters related to kidney function will be considered as secondary outcomes. A knowledge synthesis librarian developed a literature search strategy for MEDLINE (Ovid). The search strategy was then adjusted for use in Cochrane Central (Ovid), Embase (Ovid), Web of Science Core Collection (Clarivate) and CINAHL (EBSCO). Two independent reviewers will select studies for eligibility in Covidence and data extraction will be conducted using a custom MS Excel worksheet. We are planning to perform meta-analysis wherever possible using random effects model. Standardized mean difference (95% confidence interval) and risk ratio will be used to present continuous and dichotomous data, respectively. The assessment of publication bias will be performed using funnel plots and Egger’s regression test while I^2^ statistics will be used to assess heterogeneity. We are planning to perform subgroup analysis to deal with potential heterogeneity.

**Discussion:** The results of this systematic review and meta-analysis will be useful in designing effective dietary strategies for the management of CKD-related metabolic acidosis.

**Systematic review registration:** The present systematic review is registered in International Prospective Register of Systematic Reviews (PROSPERO) (https://www.crd.york.ac.uk/, registration ID: CRD42022342612).

## Background

Chronic Kidney Disease (CKD) is a progressive condition and is recognized as a leading health concern worldwide (1, 2). The condition is categorized into 5 stages based on Glomerular Filtration Rate (GFR); Early stages are asymptomatic, but can progress rapidly to kidney failure within months in some individuals (3). CKD is associated with increased risk of cardiovascular disease (CVD), kidney failure requiring renal replacement therapy, poor quality of life, and mortality (4). The global prevalence of CKD has been estimated 13.4% (5). According to a cross-sectional study, the overall estimated prevalence of stage 3 to 5 CKD in the Canadian population was 71.9 per 1000 individuals, from 2010 to 2015 (6).

Kidneys play a crucial role in maintaining acid-base balance by reabsorbing bicarbonate from filtrate and excreting acid and ammonia into urine (7). The cause of metabolic acidosis in CKD is the inability of kidneys to excrete enough acid, leading to a positive H^+^ balance and low CO2 concentrations (13). As kidney function declines metabolic acidosis becomes a common complication in people with CKD and its prevalence increases with the decline in GFR, especially when GFR levels fall below 40 ml/min/1.73m^2^ (8, 9). Although metabolic acidosis is usually mild to moderate in CKD, the condition can impair different organ systems, and contribute to increased morbidity and mortality (10, 11). Augmentation of inflammation, bone disease exacerbation, muscle wasting, and acceleration of CKD progression are among adverse effects related to metabolic acidosis (12).

Sodium bicarbonate (NaHCO3) is an alkali salt commonly used in the treatment of metabolic acidosis (14), which may improve endothelial function and muscle mass and delay kidney failure (15). However, oral alkali therapy has been related to a number of side effects including gastric discomfort and bloating, with many people having difficulty tolerating higher doses (16). Furthermore, concerns have been raised about sodium retention, volume overload and exacerbating pre-existing hypertension when sodium bicarbonate is used (17).

Daily acid load is mostly determined by the metabolism of dietary constituents into base or acid production. Sulfur containing amino acids (abundant in animal sources of protein) lead to H^+^ production while fruit and vegetables are base producing (9). Dietary H^+^ reduction is hence another strategy in the management of CKD-related metabolic acidosis, by reducing the intake of acid producing components (such as animal proteins) as opposed to adding base by increasing fruit and vegetables consumption (18).

Recent studies have reported that treating metabolic acidosis with fruit and vegetables might be as effective as alkali therapy in improving disease state by modifying HCO3 concentrations, while also reducing blood pressure and preserving eGFR with no concerns regarding adverse events (19, 20). One clinical trial has even shown that fruit and vegetables treatment not only improved metabolic acidosis, but also reduced CVD risk better than NaHCO3 (21). There is a need to integrate existing findings on the effects of adding base through fruit and vegetables consumption in the management of CKD-related metabolic acidosis. This proposed systematic review will evaluate the efficacy and safety of using fruit and vegetables to manage metabolic acidosis, as well as look at patients’ compliance and tolerability, looking to identify limitations in the current literature and set directions for future research.(23)

## Methods

### Aim

This systematic review aims to summarize findings from RCTs comparing dietary interventions focused on adding base (via fruit and vegetables consumption) with dietary interventions focusing on lowering acid load versus placebo/usual care/no treatment in the management of metabolic acidosis in outpatient adults with CKD.

This study protocol was developed using Cochrane systematic review methodology (24) and is reported according to Preferred Reporting Items for Systematic review and Meta-Analysis Protocols (PRISMA-P) guidelines (25). The protocol has been registered in International Prospective Register of Systematic Reviews (PROSPERO) (https://www.crd.york.ac.uk/, registration ID: CRD42022342612).

### Study types and eligibility criteria

The following study types will be eligible for inclusion: Randomized controlled trials, and cross-over randomized trials on adult participants (18 years of age or older), with CKD (as diagnosed using any recognized diagnostic criteria or author-defined) with eGFR between 15 and 40 ml/min/1.73 m^2^ and serum bicarbonate levels of 14-24 mEq/L. Studies will be excluded if participants are undergoing dialysis or have chronic obstructive pulmonary disease (COPD) requiring oxygen therapy.

### Intervention and control groups

Any dietary guidance or modification looking to manipulate dietary acid load will be considered as an intervention. The comparators will be usual care/ no treatment or placebo.

### Outcome measures

Our primary outcome will be change in serum bicarbonate concentrations (mEq/L). Secondary outcomes are systolic and diastolic blood pressure, heart rate, anthropometric measurements, blood urea nitrogen (BUN), creatinine, eGFR, glucose, albumin, calcium, chloride, phosphorous, potassium, sodium and HbA1c, albumin/creatinine ratio in urine samples, quality of life, reported adverse effects, mortality and KDIGO (Kidney Disease Improving Global Outcomes) criteria for acute kidney injury. This systematic review will also investigate the safety and tolerability of dietary interventions (either increasing fruit and vegetables intake or decreasing animal proteins), as well as bicarbonate therapy or placebo, in the management of metabolic acidosis in people with CKD.

### Study identification and selection

A knowledge synthesis librarian (NA) developed the literature search strategy for MEDLINE (Ovid) using a modified version of the SIGN RCT filter (26). This strategy was then peer-reviewed by a second independent librarian using the PRESS checklist (23). The final search strategy was then adjusted for use in Cochrane Central (Ovid), Embase (Ovid), Web of Science Core Collection (Clarivate) and CINAHL (EBSCO). The search strategy for this systematic review is presented in **Supplementary file 1**. Records retrieved by the search will be uploaded in Covidence systematic review management software and screened for eligibility by two independent reviewers (SM and RM). Ineligible citations will be recorded and the number and reason for exclusion will be documented at the full-text article screening stage. Disagreements on the eligibility of citations will be discussed by two reviewers with a third reviewer (DM) to adjudicate, as required. Included citations will be limited to the English language.

### Data extraction and management

Data extraction will be done using forms developed, stored and managed in MS Excel worksheet. Two reviewers will independently extract the data from all included studies. Any concerns or disagreements will be discussed and settled between the two reviewers and a third reviewer will adjudicate the issues that cannot be resolved by the two reviewers.

The following data will be extracted from the included studies:

- Study details: name of first author, year of publication, study period, country, study size, and funding source
- Study population details: type of population (age, sex distribution, comorbidities, and socioeconomic status)
- Intervention details: name, type, method of intervention, measure (amount/extent/dosage), foods provided or recommended, foods instructed to avoid, duration
- Outcome details: Data will be extracted at the end of the trial and at the longest reported follow-up.

### Risk-of-bias assessment

The risk of bias will be assessed through the Cochrane Risk of Bias Tool 2.0. (27). The tool will provide an assessment score in the forms of high, some concerns, and low risk of bias in each of the following areas: bias arising from the randomization process, bias due to deviations from intended interventions, bias due to missing outcome data, bias in measurement of the outcome, and bias in selection of the reported result. Any disagreements or issues will be discussed by the two reviewers or by involving a third reviewer if required.

### Data Analysis

We are planning to conduct meta-analysis wherever possible, using a random effects model. Continuous data will be presented as mean differences or standardized mean differences where measures of the same outcome are with different scales, with 95% confidence intervals.

Dichotomous data will be assessed and reported as a risk ratio with 95% confidence intervals. Statistical heterogeneity will be assessed between the included studies by using the I-squared (I^2^) statistic (28). Our assessment of publication bias will be completed through the use of funnel plots (29) and using Egger’s regression test (30) if there are at least 10 included trials in the meta-analysis.

We are planning to perform a prespecified subgroup analyses investigating dietary intervention method (adding base vs. reducing acid) and type of intervention (providing food vs. dietary guidance only). We will also conduct sensitivity analyses looking at diabetes comorbidity (people with diabetes vs. those without diabetes using regression against proportion of people with diabetes in trial cohort), trial type (parallel vs. crossover) sex, CKD stage, and geographical region.

## Discussion

Metabolic acidosis is of the most common consequences of CKD, with a leading role in CKD-related morbidity and mortality (31). Data from both observational and interventional studies has shown that correction of metabolic acidosis by NaHCO3 or diets rich in base producing fruit and vegetables may have the ability to slow the rate of decline in kidney function (32). Patients on Na^+^-based alkali therapy are at an increased risk of worsened hypertension or volume control, especially at lower eGFR (33). On the other hand, the usual dietary habits of developed countries often result in diets with increased production of acid equivalents (34). Therefore, dietary interventions targeting the addition of base producing fruit and vegetables may be more appropriate than alkaline therapy in the management of CKD induced metabolic acidosis. Some small single center trials have been conducted on the effects of fruit and vegetables consumption on different outcomes related to kidney function, in different stages of CKD. Some review articles have included effects of dietary interventions in the management of CKD (34, 35) which have provided general information in the topic. To the best of our knowledge, our study will be the first to systematically review data from randomized studies aiming at evaluating the effects of adding dietary base equivalents via fruit and vegetables consumption in the management of CKD-related metabolic acidosis. Our study is not obstacle-free. The number of studies meeting our criteria might be few and heterogeneity might arise due to different settings and clinical stages. Nevertheless, we have prepared a plan for prespecified subgroup analysis based on dietary interventions (adding base vs. reducing acid), type of intervention (providing food vs. recommendation only), sex, diabetes comorbidity, CKD stages and region. The findings of this study will be useful to guide clinical practice regarding the use of base producing fruit and vegetables as an alternative option to oral alkali therapy. Furthermore, this study will find limitations of the current literature and inform the design of future research in this area

## Data Availability

All data produced in the present study are available upon reasonable request to the authors

## Abbreviations

BUN: Blood Urea Nitrogen
CKD: Chronic Kidney Disease
COPD: Chronic Obstructive Pulmonary Disease
CVD: Cardiovascular Disease
GFR: Glomerular Filtration Rate
KDIGO: Kidney Disease Improving Global Outcomes
PRISMA-P: Preferred Reporting Items for Systematic review and Meta-Analysis Protocols
PROSPERO: Prospective Register of Systematic Reviews
RCT: Randomized Clinical Trial

## Supplementary information

Supplementary file 1: search strategy Supplementary file 2: PRISMA-P 2015 checklist

## Acknowledgement

We would like to thank Tyler Ostapyk for conducting the PRESS review of our search strategy.

## Authors contribution

SM drafted the review protocol manuscript. NA is a health sciences librarian who developed the search strategy based on input from DM, RM, AMAS, RR NT and SM. AAB and RB provided content expertise in knowledge synthesis and meta-analysis. NT provided content expertise in chronic kidney disease. RM and DM are content experts in nutritional interventions. Each author reviewed and made substantial contributions to this manuscript. All authors have read and approved the final manuscript.

## Availability of data and material

The Endnote database, as well as final extraction sheets and data regarding risk of bias assessment will be available upon request.

## Ethics approval and consent to participate

Not applicable

## Consent for publication

Not applicable

