## Supplementary Table 1 for "Effects of dietary interventions focused on adding base in the management of metabolic acidosis in adults with chronic kidney disease: A protocol for systematic review and meta-analysis of randomized clinical trials"

Supplementary file 1. Search Strategy

| Database | Search strategy | Results | Summary |
|---|---|---|---|
| Medline (Ovid) | 1. exp renal insufficiency, chronic/ or uremia/<br>2. (((chronic or endstage or end-stage or advanced or insufficien*) adj2 (kidney or renal)) or ckd or ckf or crf or crd or esrf or eskf or esrd or eskd or ur?emi* or frasier or gfr or egfr or estimated glomerular).ti,ab,kf<br>3. or/1-2<br>4. acidosis/ or acidosis, lactic/ or acidosis, renal tubular/ or exp ketosis/<br>5. (((acid or base or acidbase) adj2 (balanc* or imbalanc*)) or (bicarb* adj2 (serum* or blood* or level*)) or total co2 or ketoacid* or keto acid* or ketosis or lactacidos* or aceton?emi* or keton?emi* or acetonuri* or ammoniagenesis or rta or rtas).ti,ab,kf<br>6. ((metaboli* or nonrespiratory or non-respiratory or renal or kidney or tubul* or lactic or lactate) adj3 acido*).ti,ab,kf<br>7. or/4-6<br>8. exp vegetarians/ or exp diet therapy/ or exp "food and beverages"/ or exp diet/ or eating/<br>9. dh.fs<br>10. (diet* or nutrition* or food or foods or eat or eating or nutr#ceutical* or ketoanalogue* or keto or fruit* or vegetable* or vegetarian* or lactovegetarian* or vegan* or mediterranean or plant or plants or juice* or meat or meats or legume* or nut or nuts or seed or seeds or salad*).ti,ab,kf<br>11. ((reduc* or low or lower* or restrict* or free or intak* or decreas*) adj2 (protein*)).ti,ab,kf<br>12. or/8-11<br>13. randomized controlled trial/ or random allocation/ or double blind method/ or single blind method/ or clinical trial/<br>14. ("clinical trial, phase I" or "clinical trial, phase ii" or "clinical trial, phase iii" or "clinical trial, phase iv" or controlled clinical trial or randomized controlled trial or multicenter study or clinical trial).pt<br>15. exp clinical trials as topic/<br>16. (clinical adj trial*).tw<br>17. ((singl* or doub* or treb* or tripl*) adj (blind* or mask*)).tw | 143329<br>287111<br>335678<br>32145<br>31455<br>22873<br>65726<br>1687066<br>55265<br>2131345<br>105148<br>3234574<br>999192<br>1132028<br>375048<br>439796<br>189359 | 185 |

| | | | |
|---|---|---|---|
| | 18. placebos/<br>19. (placebo*).tw<br>20. (random* adj2 allocat*).tw<br>21. or/13-20<br>22. exp animals/ not humans/<br>23. 21 not 22<br>24. 3 and 7 and 12 and 23 | 35916<br>236694<br>40765<br>1834974<br>5020785<br>1735792<br>185 | |
| Embase (Ovid) | 1. exp chronic kidney failure/ or end stage renal disease/ or frasier syndrome/ or mild renal impairment/ or moderate renal impairment/ or severe renal impairment/ or uremia/<br>2.(((chronic or endstage or end-stage or advanced or insufficien*) adj2 (kidney or renal)) or ckd or ckf or crf or crd or esrf or eskf or esrd or eskd or ur?emi* or frasier or gfr or egfr or estimated glomerular).ti,ab,kw<br>3. or/1-2<br>4. acidosis/ or metabolic acidosis/ or diabetic ketoacidosis/ or hyperchloremic acidosis/ or ketoacidosis/ or kidney tubule acidosis/ or lactic acidosis/<br>5. (((acid or base or acidbase) adj2 (balanc* or imbalanc*)) or (bicarb* adj2 (serum* or blood* or level*)) or total co2 or ketoacid* or keto acid* or ketosis or lactacidos* or aceton?emi* or keton?emi* or acetonuri* or ammoniagenesis or rta or rtas).ti,ab,kw<br>6. ((metaboli* or nonrespiratory or non-respiratory or renal or kidney or tubul* or lactic or lactate) adj3 acido*).ti,ab,kw<br>7. or/4-6<br>8. exp vegetarian/ or vegan/ or exp diet therapy/ or exp diet/ or exp dietary intake/ or dietary pattern/ or exp food/ or exp food intake/ or meal/<br>9. (diet* or nutrition* or food or foods or eat or eating or nutr#ceutical* or ketoanalogue* or keto or fruit* or vegetable* or vegetarian* or lactovegetarian* or vegan* or mediterranean or plant or plants or juice* or meat or meats or legume* or nut or nuts or seed or seeds or salad*).ti,ab,kw<br>10. ((reduc* or low or lower* or restrict* or free or intak* or decreas*) adj2 (protein*)).ti,ab,kw<br>11. or/8-10<br>12. clinical trial/ or exp randomized controlled trial/ or controlled clinical trial/ or multicenter study/ or phase 3 clinical trial/ or phase 4 clinical trial/ or exp | 189936<br><br><br>446796<br><br><br>504765<br>78329<br><br>41103<br><br><br><br>32085<br><br>107661<br>2154326<br><br>2458489<br><br><br><br>131810<br><br>3554724<br><br>1754843 | 407 |

| | | | |
|---|---|---|---|
| | randomization/ or single blind procedure/ or double blind procedure/ or triple blind procedure/ or crossover procedure/<br>13. exp "clinical trial (topic)"/<br>14. (clinical adj trial*).tw<br>15. ((singl* or doub* or treb* or tripl*) adj (blind* or mask*)).tw<br>16. placebo/<br>17. (placebo*).tw<br>18. (random* adj2 allocat*).tw<br>19. or/12-18<br>20. (exp animal/ or nonhuman/) not exp human/<br>21. 19 not 20<br>22. 3 and 7 and 11 and 21 | <br><br>393660<br>626958<br>264448<br>381528<br>344192<br>50249<br>2576241<br>6837264<br>2469463<br>407 | |
| Cochrane Central (Ovid) | 1. exp renal insufficiency, chronic/ or uremia/<br>2. (((chronic or endstage or end-stage or advanced or insufficien*) adj2 (kidney or renal)) or ckd or ckf or crf or crd or esrf or eskf or esrd or eskd or ur?emi* or frasier or gfr or egfr or estimated glomerular).ti,ab,kw<br>3.or/1-2<br>4. acidosis/ or acidosis, lactic/ or acidosis, renal tubular/ or exp ketosis/<br>5. (((acid or base or acidbase) adj2 (balanc* or imbalanc*)) or (bicarb* adj2 (serum* or blood* or level*)) or total co2 or ketoacid* or keto acid* or ketosis or lactacidos* or aceton?emi* or keton?emi* or acetonuri* or ammoniagenesis or rta or rtas).ti,ab,kw<br>6. ((metaboli* or nonrespiratory or non-respiratory or renal or kidney or tubul* or lactic or lactate) adj3 acido*).ti,ab,kw<br>7. or/4-6<br>8. exp vegetarians/ or exp diet therapy/ or exp "food and beverages"/ or exp diet/ or eating/<br>9. dh.fs<br>10. (diet* or nutrition* or food or foods or eat or eating or nutr#ceutical* or ketoanalogue* or keto or fruit* or vegetable* or vegetarian* or lactovegetarian* or vegan* or mediterranean or plant or plants or juice* or meat or meats or legume* or nut or nuts or seed or seeds or salad*).ti,ab,kw<br>11. ((reduc* or low or lower* or restrict* or free or intak* or decreas*) adj2 (protein*)).ti,ab,kw<br>12. or/8-11 | 7773<br>36681<br><br><br>38893<br>676<br>2963<br><br><br><br>1513<br><br>4344<br>68202<br><br>8482<br>159174<br><br><br><br>8996<br>196721<br>236 | 236 |

| | | | |
|---|---|---|---|
| | 13. 3 and 7 and 12 | | |
| CINAHL with full text (EBSCO) | 1. (MH "renal insufficiency, chronic+") or (MH uremia)<br>2. (((chronic or endstage or "end stage" or advanced or insufficien*) N2 (kidney or renal)) or ckd or ckf or crf or crd or esrf or eskf or esrd or eskd or ur#emi* or frasier or gfr or egfr or "estimated glomerular")<br>3. S1 or S2<br>4. (MH acidosis) or (MH "acidosis, lactic") or (MH "diabetic ketoacidosis")<br>5. (((acid or base or acidbase) N2 (balanc* or imbalanc*)) or (bicarb* N2 (serum* or blood* or level*)) or "total co2" or ketoacid* or (keto N1 acid*) or ketosis or lactacidos* or aceton#emi* or keton#emi* or acetonuri* or ammoniagenesis or rta or rtas)<br>6. ((metaboli* or nonrespiratory or "non respiratory" or renal or kidney or tubul* or lactic or lactate) N3 acido*)<br>7. S4 or S5 or S6<br>8. (MH diet+) or (MH "diet therapy+") or (MH "food and beverages+") or (MH eating)<br>9. MW "DH"<br>10. (diet* or nutrition* or food or foods or eat or eating or nutr?ceutical* or ketoanalogue* or keto or fruit* or vegetable* or vegetarian* or lactovegetarian* or vegan* or mediterranean or plant or plants or juice* or meat or meats or legume* or nut or nuts or seed or seeds or salad*)<br>11. ((reduc* or low or lower* or restrict* or free or intak* or decreas*) N2 (protein*))<br>12. S8 or S9 or S10 or S11<br>13. (MH "clinical trials+")<br>14. PT clinical trial<br>15. TX (clinical N1 trial*)<br>16. TX ((singl* or doub* or treb* or tripl*) N1 (blind* or mask*))<br>17. (MH "random assignment") or (MH placebos) or (MH "quantitative studies")<br>18. TX(placebo*)<br>19. TX(random* N2 allocat*)<br>20. S13 or S14 or S15 or S16 or S17 or S18 or S19 | 32162<br>85954<br><br>85954<br>6775<br>14093<br><br>5691<br>18502<br>307046<br>23805<br>602205<br><br>15329<br>665662<br>338222<br>111341<br>379547<br>1244043<br>116788<br>105701<br>19588<br>1653788 | 91 |

| | | | |
|---|---|---|---|
| | 21. (MH vertebrates+) not (MH human)<br>22. S20 not S21<br>23. S3 and S7 and S12 and S22 | 213128<br>1626191<br>91 | |
| Web of Science Core Collection | 1. TS=(((chronic or endstage or "end stage" or advanced or insufficien*) NEAR/2 (kidney or renal)) or ckd or ckf or crf or crd or esrf or eskf or esrd or eskd or ur$emi* or frasier or gfr or egfr or "estimated glomerular")<br>2. TS=(((acid or base or acidbase) NEAR/2 (balanc* or imbalanc*)) or (bicarb* NEAR/2 (serum* or blood* or level*)) or "total co2" or ketoacid* or (keto NEAR/1 acid*) or ketosis or lactacidos* or aceton$emi* or keton$emi* or acetonuri* or ammoniagenesis or rta or rtas)<br>3. TS=((metaboli* or nonrespiratory or "non respiratory" or renal or kidney or tubul* or lactic or lactate) NEAR/3 acido*)<br>4. #2 or #3<br>5. TS=(diet* or nutrition* or food or foods or eat or eating or nutr?ceutical* or ketoanalogue* or keto or fruit* or vegetable* or vegetarian* or lactovegetarian* or vegan* or mediterranean or plant or plants or juice* or meat or meats or legume* or nut or nuts or seed or seeds or salad*)<br>6. TS=((reduc* or low or lower* or restrict* or free or intak* or decreas*) NEAR/2 (protein*))<br>7. #5 or #6<br>8. TS=(clinical NEAR/1 trial*)<br>9. TS=((singl* or doub* or treb* or tripl*) NEAR/1 (blind* or mask*))<br>10. TS=(placebo*)<br>11. TS=(random* NEAR/2 allocat*)<br>12. #8 or #9 or #10 or #11<br>13. #1 and #4 and #7 and #12 | 345155<br>66381<br>26278<br>89452<br>4665420<br>174932<br>4783839<br>515579<br>343214<br>279718<br>41683<br>923741<br>115 | 115 |
